# Association between aspirin dose and outcomes in patients with acute Kawasaki disease

**DOI:** 10.1101/2023.02.13.23285893

**Authors:** Takanori Suzuki, Nobuaki Michihata, Yohei Hashimoto, Tetsushi Yoshikawa, Kazuyoshi Saito, Hiroki Matsui, Kiyohide Fushimi, Hideo Yasunaga

**Author notes:** **Correspondence to:** Nobuaki Michihata, MD, MPH, PhD, Department of Health Services Research, Graduate School of Medicine, The University of Tokyo, 7-3-1 Hongo, Bunkyo-ku, Tokyo 113-0033, Japan. **Open Access:** We do not want open access. **Data sharing statement:** The datasets analyzed during the current study are not publicly available due to contracts with the hospitals providing data to the database. **Author contributions:** Takanori Suzuki wrote the first draft of the manuscript. Nobuaki Michihata conceptualized and designed the study, drafted the initial manuscript, and reviewed and revised the manuscript. Kazuyoshi Saito, Tetsushi Yoshikawa, Hiroki Matsui, Kiyohide Fushimi, and Hideo Yasunaga conceptualized and designed the study, and coordinated and critically reviewed the manuscript for important intellectual content. All authors approved the final manuscript as submitted and agree to be accountable for all aspects of the work.

## Abstract

**Background:** The most effective dosage of aspirin to prevent coronary artery abnormalities in patients with acute Kawasaki disease remains unknown. Using a Japanese national inpatient database, this study aimed to identify the appropriate dose of aspirin to be prescribed to patients with acute Kawasaki disease.

**Method:** We used the Diagnostic Procedure Combination database to identify patients with Kawasaki disease treated with intravenous immunoglobulin between 2010 and 2021.The outcomes included the occurrence of coronary artery abnormalities and intravenous immunoglobulin resistance, length of stay, and medical costs. Restricted cubic spline functions were performed to examine the association between aspirin dose and the outcomes.

**Results:** Data of 82109 patients were extracted from the database. Non-linear associations were observed between aspirin dose and the outcomes. In comparison with an aspirin dose of 30 mg/kg/day, the odds ratio (95% confidence interval) for coronary artery abnormalities was 1.40 (1.13–1.75) at 5 mg/kg/day. An aspirin dose of ≥30 mg/kg/day did not significantly change the odds ratio for coronary artery abnormalities. Compared with a dose of 30 mg/kg/day, the odds ratio (95% confidence interval) for intravenous immunoglobulin resistance was 0.87 (0.76–1.00) at 5 mg/kg/day and 0.59 (0.36–0.98) at 80 mg/kg/day. An increase in aspirin dose was associated with a shorter length of stay and lower medical costs.

**Conclusions:** Low-dose aspirin may increase the risk of coronary artery abnormalities in patients with acute Kawasaki disease; however, increasing aspirin doses beyond the standard doses may not be associated with a reduction in coronary artery abnormalities. High-dose aspirin showed the potential to reduce hospital stay and medical costs without increasing complications.

**Article Summary:** The study showed by restricted cubic spline that the dose of aspirin was no significant association between aspirin escalation and CAAs in patients with acute KD.

**What’s Known on This Subject:** Aspirin is standard treatments used with IVIG of acute Kawasaki Disease (KD), Few studies have shown the most effective dosage of aspirin to to prevent CAAs.

**What This Study Adds:** The dose of aspirin was no significant association between aspirin escalation and CAAs in patients with acute KD.

## Introduction

Kawasaki disease (KD) represents an acute form of systemic vasculitis, which most commonly leads to acquired heart disease in children in most industrialized countries.^1^ The standard treatment for patients with acute KD is the administration of aspirin and intravenous immunoglobulin (IVIG). As for the aspirin dose for patients with acute KD, the American Heart Association guidelines for KD recommend 80–100 mg/kg/day, while the Japanese Society of Pediatric Cardiology and Cardiac Surgery guidelines for KD recommend 30–50 mg/kg/day in the acute KD.^2,3^ However, various trials have investigated the effectiveness of different aspirin doses for patients with acute KD. Recent small-scale studies have reported no difference in coronary artery abnormalities (CAAs) outcomes with aspirin doses as low as 3–5 mg/kg/day or as high as 100 mg/kg/day.^4–9^ Another study found that high doses of aspirin enhanced the production of tumor necrosis factor-alpha (TNF-α), which is related to the development of CAAs.^11^ Thus, the results of previous studies are controversial, and the most appropriate dose of aspirin remains unknown. Conversely, it was recently reported that high-dose aspirin may have a negative effect on the initial IVIG therapy during the acute febrile phase of KD.^10^ This study aimed to examine the association between aspirin dose and outcomes in patients with acute KD, using a Japanese national inpatient database.

## Methods

### Data source

For this retrospective cohort study, we used data from the Diagnosis Procedure Combination database. Over 80 academic hospitals are obliged to participate in the database; however, the participation of community hospitals in the database is voluntary. Data for about 8 million hospitalized patients of all ages are included every year. It is almost equivalent to 50% of the total acute-care hospitalizations in Japan. The database includes the following information: unique identifiers of hospitals, patient baseline characteristics, and diagnosis at admission, comorbidities at admission, and complications after admission recorded with text data in Japanese and International Classification of Diseases, Tenth Revision (ICD-10) codes; medical procedures and treatments, including drug administrations, devices use, and surgical and nonsurgical procedures, length of stay, discharge status, and medical cost during hospitalization. Attending physicians must record diagnoses and comorbidities with reference to medical records. The current study was approved by the Institutional Review Board of The University of Tokyo (approval number: 3501-(5) (May 19th, 2021). The requirement for informed consent was waived due to the anonymous nature of the data.

### Participants

We identified patients diagnosed with KD (ICD-10 code: M303) between July 2010 and March 2021. We included patients who were treated with initial IVIG treatment. We further checked the Japanese text describing the detailed diagnoses in each case to include patients with atypical KD and exclude patients with a ‘suspected’ diagnosis of KD. We excluded patients who weighed <3 kg, >60 kg or had missing data. Patients who used flurbiprofen were excluded from our study. The initial aspirin dose was defined as the dose used at the same time as the initial IVIG in the acute phase of KD. We also excluded initial aspirin dose <3 mg/kg/day or initial aspirin dose >90 mg/kg/day. We classified the study population according to the aspirin dose at the initial treatment of IVIG: low-dose group, aspirin <30 mg/kg/day; standard-dose group, 30–50 mg/kg/day; and high-dose group, >50 mg/kg/day.

### Outcomes

The primary outcome was the occurrence of CAAs upon discharge. CAAs were identified with a recorded diagnosis of CAAs (ICD-10 code: I25.4) and/or text data of CAAs in the Japanese language. The secondary outcomes were IVIG resistance, length of stay, medical costs, and adverse events. We converted 1 United States dollar (USD) to 100 Japanese yen. IVIG resistance was defined as the use of IVIG at a total dose of ≥4.0 g/kg and/or a combination of any steroid, infliximab, cyclosporine, and/or plasma exchange that was not administered during the initial IVIG treatment. The medical costs included costs of surgery, drugs, laboratory tests, and other inpatient services, including food expenses. We also examined adverse events after admission occurring with aspirin, including Reye syndrome (ICD-10 code: G937), bleeding from the upper gastrointestinal tract (ICD-10 code: K922), anemia (ICD-10 code: D5, D60-4), gastric ulcer (ICD-10 code: K25-6), and liver function abnormality (ICD-10 code: K719, K769) /or text data of adverse events in the Japanese language.

### Covariates

The baseline characteristics included sex, age, weight, height, hospital days of illness at initial IVIG, use of 10% IVIG, initial steroid use, initial cyclosporine use, type of hospital, complex chronic conditions,^12^ Japan Coma Scale at admission, transportation by ambulance, transportation from other hospitals, and hospital volume. Initial steroid use and initial cyclosporine use were defined as those concurrent with initial IVIG use. For the hospital type, academic hospitals were defined as university hospitals and related hospitals. The Japan Coma Scale score at admission was categorized into two groups: alert and not alert. Japan Coma Scale assessment was previously clarified to be well associated with the Glasgow Coma Scale assessment.^13^ Hospital volume was defined as the number of all patients with KD at each hospital every year. Hospital volume was divided into tertiles so that the number of patients in each group was almost equal.

### Statistical analysis

Categorical variables were presented as numbers and percentages and were compared using the Fisher exact test. Continuous variables were presented as means and standard deviations (SD) or medians and interquartile ranges (IQR). Non-normally distributed variables (length of stay and medical costs) were compared among the three groups using the Kruskal–Wallis test. The sensitivity analysis of CAAs was limited to those who had a diagnosis of CAAs and were using anticoagulants, such as warfarin or clopidogrel, or were undergoing cardiac catheterization.

### Restricted cubic spline functions

Studies regarding the relationship between aspirin dose and outcomes converted continuous measures of aspirin into categorical variables. However, this may have resulted in a loss of information and a decrease in statistical power.^14^ Therefore, we used multivariable regression models with restricted cubic spline functions to assess the potential non-linear association of aspirin dose with CAAs, IVIG resistance, length of stay, and medical cost. Multivariable logistic regression models were applied for the occurrence of CAAs and IVIG resistance, and multiple regression models were applied for length of stay and medical cost with adjustment for age, sex, body weight, body height, hospital days of illness at the initial IVIG treatment, use of 10% IVIG, initial steroid use, initial cyclosporine use, type of hospital, complex chronic conditions, transportation by ambulance, Japan Coma Scale at admission, transportation from another hospital, fiscal year, and hospital volume. We created restricted cubic splines with five knots at pre-specified locations according to the percentiles of the distribution of aspirin doses (10, 30, 40, 50, and 70 mg/kg/day). Tests for overall and non-linear associations were performed using the χ^2^ test.^15^ A two-sided value of *P*<.05 was considered significant. Statistical analyses were performed using the Stata software 17 (StataCorp LP, TX, USA).

## Results

Patient characteristics are shown in Table 1. Initially, 82,109 patients with acute KD were identified using the inclusion criteria (Figure 1). The mean age was 2.1 years (SD, 2.0), and the mean body weight was 12.3 kg (SD, 4.7). Of them, 47,277 (57.6%) were male. The median total amount of aspirin was 30.1 mg/kg/day (IQR, 29.4–32.3). Regarding the aspirin dose, the low-dose group (n=30,439) had a median of 28.9 mg/kg/day (IQR, 27.5– 29.5), the standard-dose group (n=49,646) had a median of 31.1 mg/kg/day (IQR, 30.2– 36.4), and the high-dose group (n=2,024) had a median of 52.1 mg/kg/day (IQR, 50.8– 58.5) (*P*<.001). Moreover, the high-dose group was associated with younger age, smaller weight, lower height, longer hospital days of illness, 10% IVIG and initial steroid, initial cyclosporine, academic hospital, the score of complex chronic conditions, and transport ambulance. CAAs and IVIG resistance occurred in 1817 (2.2%) and 15717 (19.1%) patients, respectively. The median (IQR) of length of stay and medical costs was 10 days (8.0–12.0) and 6454 USD (5299–8169), respectively.

**Table 1.** Patients Characteristics

| Variables | Total<br>(N = 82109) | Low-dose group<br>(N =30349) | Standard-dose group<br>(N=49646) | High-dose group<br>(N=2024) | P value |
| --- | --- | --- | --- | --- | --- |
| Age (years), mean (SD) | 2.1 (2.0) | 2.2 (2.1) | 2.1 (2.0) | 2.1 (2.1) | <.001 |
| Male (%) | 47277 (57.6%) | 17553 (57.7%) | 28561 (57.5%) | 1163 (57.5%) | .92 |
| Weight (kg), mean (SD) | 12.3 (4.7) | 12.5 (5.0) | 12.1 (4.4) | 11.2 (4.8) | <.001 |
| Height (cm), mean (SD) | 77.5 (32.0) | 78.4 (30.8) | 76.9 (32.8) | 76.4 (29.2) | <.001 |
| Hospital days of illness at initial IVIG,<br>median (IQR) | 2 (1.0-2.0) | 2 (1.0-2.0) | 2 (1.0-2.0) | 2 (1.0-3.0) | .002 |
| Total amount of initial aspirin (mg/kg/day),<br>median (IQR) | 30.1 (29.4-32.3) | 28.9 (27.5-29.5) | 31.1 (30.2-36.4) | 52.1 (50.8-58.5) | <.001 |
| Use of 10% IVIG | 9266 (11.5%) | 3351 (11.2%) | 5626 (11.5%) | 289 (14.5%) | <.001 |
| Initial steroid use | 8451 (10.3%) | 3445 (11.3%) | 4830 (9.7%) | 176 (8.7%) | <.001 |
| Initial cyclosporine use | 233 (0.3%) | 99 (0.3%) | 120 (0.2%) | 14 (0.7%) | <.001 |
| Adverse event after admission |  |  |  |  |  |
| Reye syndrome | 0 | 0 | 0 | 0 |  |
| Bleeding from the upper gastrointestinal<br>tract | 340 (0.4%) | 135 (0.4%) | 200 (0.4%) | 5 (0.2%) | .34 |
| Anemia | 735 (0.9%) | 278 (0.9%) | 432 (0.9%) | 25 (1.2%) | .21 |
| Gastric ulcer | 762 (0.9%) | 281 (0.9%) | 463 (0.9%) | 18 (0.9%) | .97 |
| Liver function abnormality | 216 (0.3%) | 78 (0.3%) | 135 (0.3%) | 3 (0.1%) | .54 |
| Type of hospital |  |  |  |  | <.001 |
| Academic hospital | 15092 (20.6%) | 6111 (22.2%) | 8405 (19.1%) | 576 (32.3%) |  |
| Complex Chronic Conditions |  |  |  |  |  |
| 0-1 | 78846 (96.0%) | 29201 (95.9%) | 47722 (96.1%) | 1923 (95.0%) | .024 |
| 2 ≤ | 3263 (4.0%) | 1238 (4.1%) | 1924 (3.9%) | 101 (5.0%) |  |
| Transportation by ambulance | 1928 (2.3%) | 758 (2.5%) | 1100 (2.2%) | 70 (3.5%) | <.001 |
| Japan coma scale at admission |  |  |  |  | .464 |
| Alert | 81454 (99.2%) | 30181 (99.2%) | 49265 (99.2%) | 2008 (99.2%) |  |
| Not Alert | 655 (0.8%) | 258 (0.8%) | 381 (0.8%) | 16 (0.8%) |  |
| Transported from another hospital | 63212 (77.0%) | 23869 (78.4%) | 37865 (76.3%) | 1478 (73.0%) | <.001 |
| Hospital volume |  |  |  |  |  |
| Low (1-29.0) | 25255 (30.8%) | 8876 (29.2%) | 15871 (32.0%) | 508 (25.1%) | <.001 |
| Middle (29.0-53.5) | 28233 (34.4%) | 10678 (35.1%) | 16971 (34.2%) | 584 (28.9%) |  |
| High (≥ 53.6) | 28621 (34.9%) | 10885 (35.8%) | 16804 (33.8%) | 932 (46.0%) |  |

IQR: interquartile range; IVIG: intravenous immunoglobulin; SD: standard deviation

**Figure 1.**
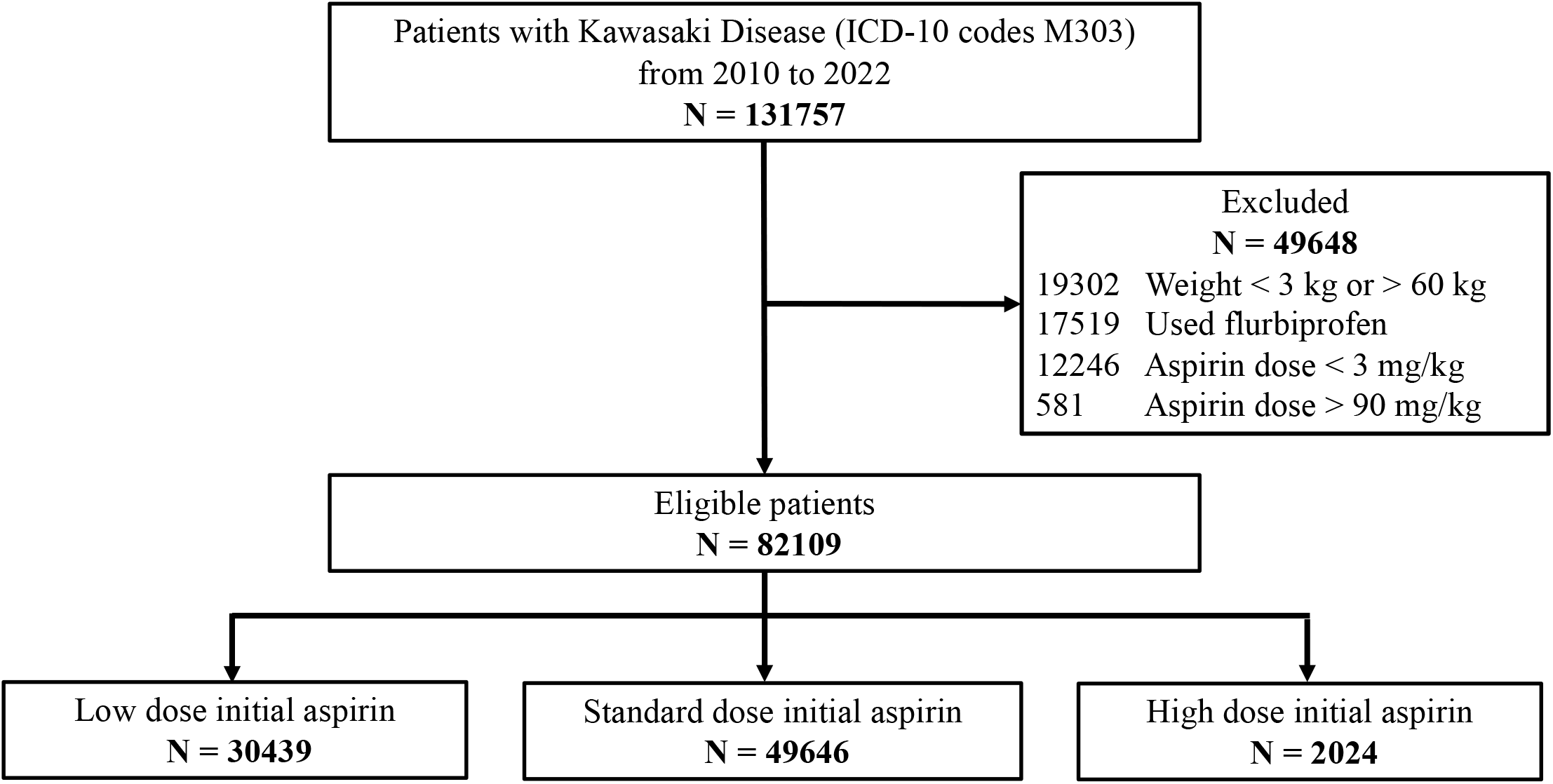
Flow chart for the study cohort ICD-10: International Classification of Diseases, Tenth Revision; IVIG: intravenous immunoglobulin; KD: Kawasaki disease

Compared with the low- and high-dose groups, the standard-dose group showed lower proportions of CAAs (2.0%, *P*<.001), shorter length of stay (median, 10 days; *P*<.001), and lower medical costs (median, 6440 USD; *P*<.001) (Table 2). No patients had Reye syndrome. There were no significant differences between the groups in the proportion of other adverse events (bleeding from the upper gastrointestinal tract, anemia, gastric ulcer, and liver function abnormality) (Table 1).

**Table 2.** Comparison of outcomes among the three groups with different doses of Aspirin

| Variables | Total<br>(N =82109) | Low-dose group<br>(N =30439) | Standard-dose group<br>(N=49646) | High-dose group<br>(N=2024) | *P* value |
| --- | --- | --- | --- | --- | --- |
| Proportion of CAAs (%) | 1817 (2.2%) | 741 (2.4%) | 1014 (2.0%) | 62 (3.1%) | <.001 |
| Proportion of IVIG resistance (%) | 15717 (19.1%) | 5403 (17.8%) | 9765 (19.7%) | 549 (27.1%) | <.001 |
| Length of stay (day) | 10.0 (8.0-12.0) | 10.0 (8.0-12.0) | 10.0 (8.0-12.0) | 10.0 (8.0-13.0) | <.001 |
| Medical cost (USD) | 6454 (5299-8169) | 6461 (5299-8184) | 6440 (5292-8139) | 6806 (5470-8895) | <.001 |
| Proportion of CAAs (sensitivity analysis **) (%) | 527 (0.6%) | 227 (0.7%) | 281 (0.6%) | 19 (0.9%) | .002 |

CAAs: coronary artery abnormalities; IVIG: intravenous immunoglobulin; USD: United States dollars
\* All data are presented as medians (interquartile ranges).
\*\* CAA in the sensitivity analysis was defined as a diagnosis of CAA (the main analysis was diagnosis only) and drug therapy (warfarin or clopidogrel) and/or cardiac catheterization.

In the sensitivity analysis, CAAs were confirmed in 527 patients (0.6%) among the study participants. Similar to the main analysis, the standard dose group showed the lowest proportion of CAAs (0.6%, *P*<.001).

Regarding restricted cubic spline functions, tests for overall association and non-linear association showed significant non-linear relationships between aspirin dose and the outcomes. In comparison with an aspirin dose of 30 mg/kg/day, the odds ratio (95% confidence interval [CI]) for CAA was 1.40 (1.13–1.75) at 5 mg/kg/day. For aspirin doses of 30 mg/kg/day or higher, the 95% CIs for the occurrence of CAA were almost across 1 and not significantly different when compared with the values observed at 30 mg/kg/day (Figure 2).

**Figure 2.**
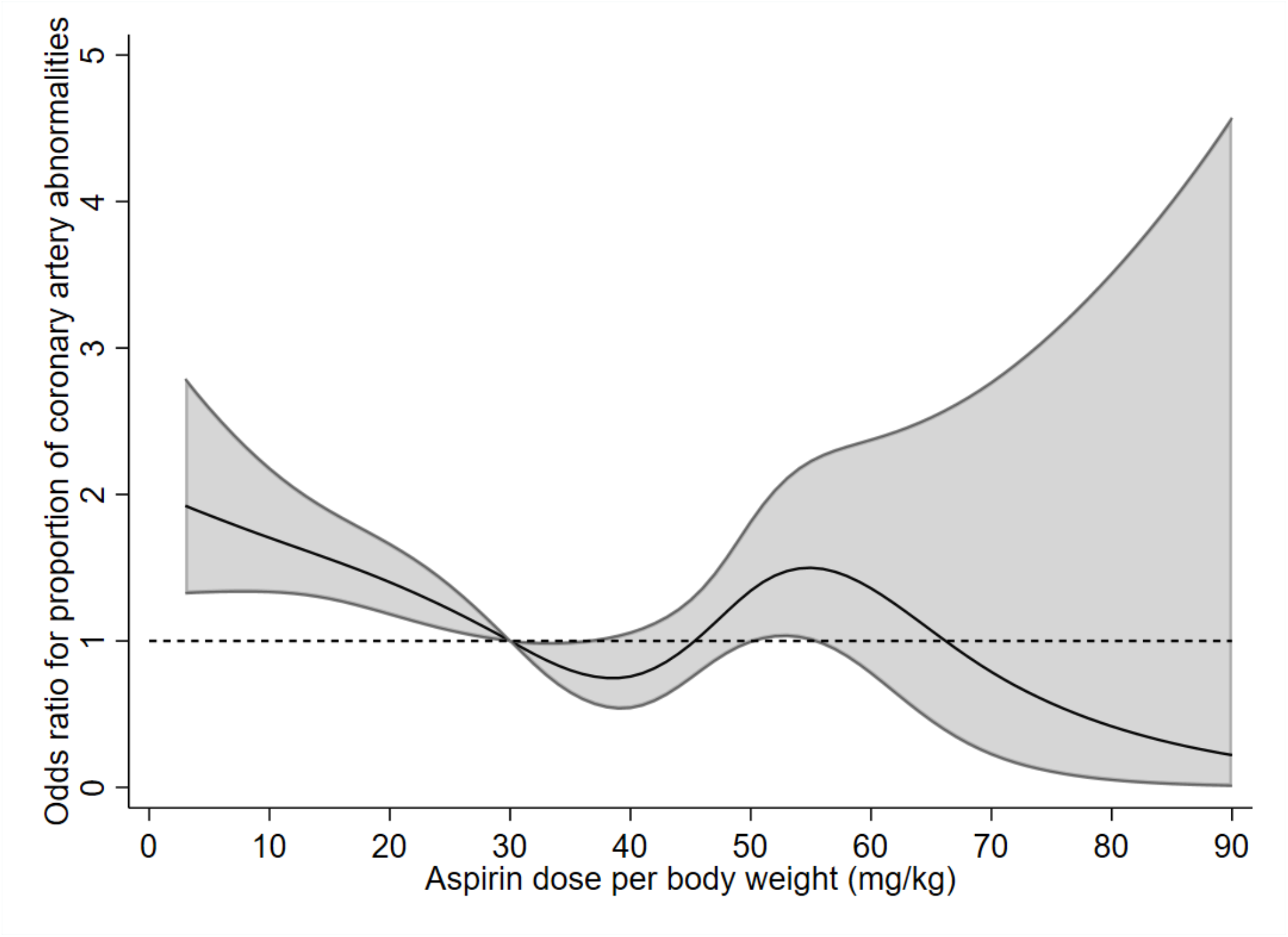
Restricted cubic spline for coronary artery abnormalities The dose–response relationship between aspirin dose and proportion of coronary artery abnormalities was adjusted by analysis using a restricted cubic spline with five knots (Aspirin, 10, 30, 40, 50, and 70 mg/kg/day). The graphs show the estimated values along with 95% CI. IVIG: intravenous immunoglobulin; CI: confidence interval

The odds ratio (95% CI) for IVIG resistance was 0.87 (0.76–1.0) at the aspirin dose of 5 mg/kg/day and 0.59 (0.36–0.98) at 80 mg/kg/day when compared with 30 mg/kg/day (Figure 3). The difference (95% CI) in the length of stay was 0.33 (0.15–0.5) days at 5 mg/kg/day and 2.32 (1.61–3.03) days at 80 mg/kg/day with reference to 30 mg/kg/day (Figure 4). The difference in medical costs was 55.8 (42.8–154.4) USD at 5 mg/kg/day and −2,307 (−2,707-−1,907) USD at 80 mg/kg/day when compared with the values observed at 30 mg/kg/day (Figure 5).

**Figure 3.**
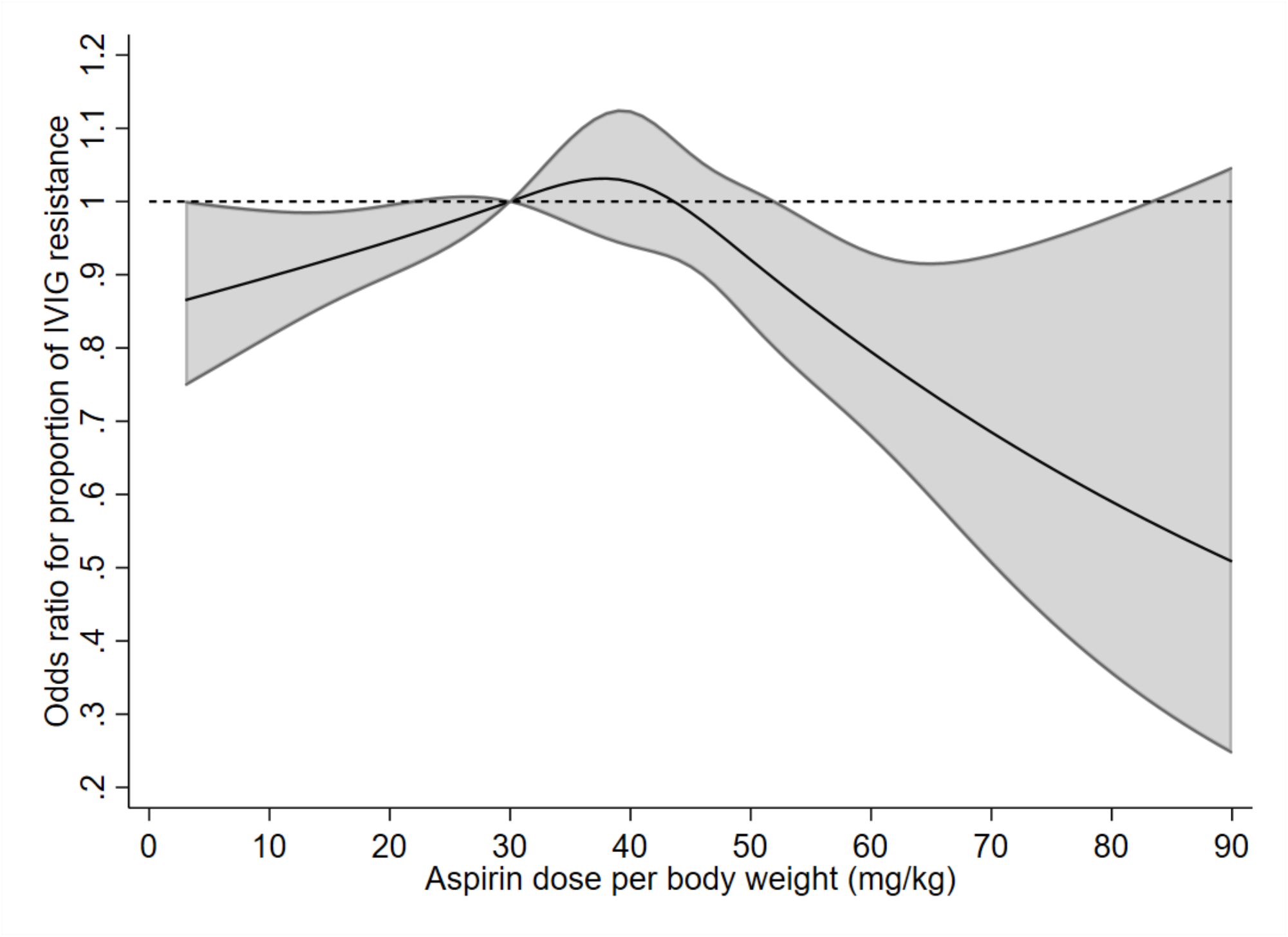
Restricted cubic spline for intravenous immunoglobulin resistance The dose–response relationship between aspirin dose and proportion of intravenous immunoglobulin resistance was adjusted by analysis using a restricted cubic spline with five knots (Aspirin, 10, 30, 40, 50, and 70 mg/kg/day). The graphs show the estimated values along with 95% CI. IVIG: intravenous immunoglobulin; CI: confidence interval

**Figure 4.**
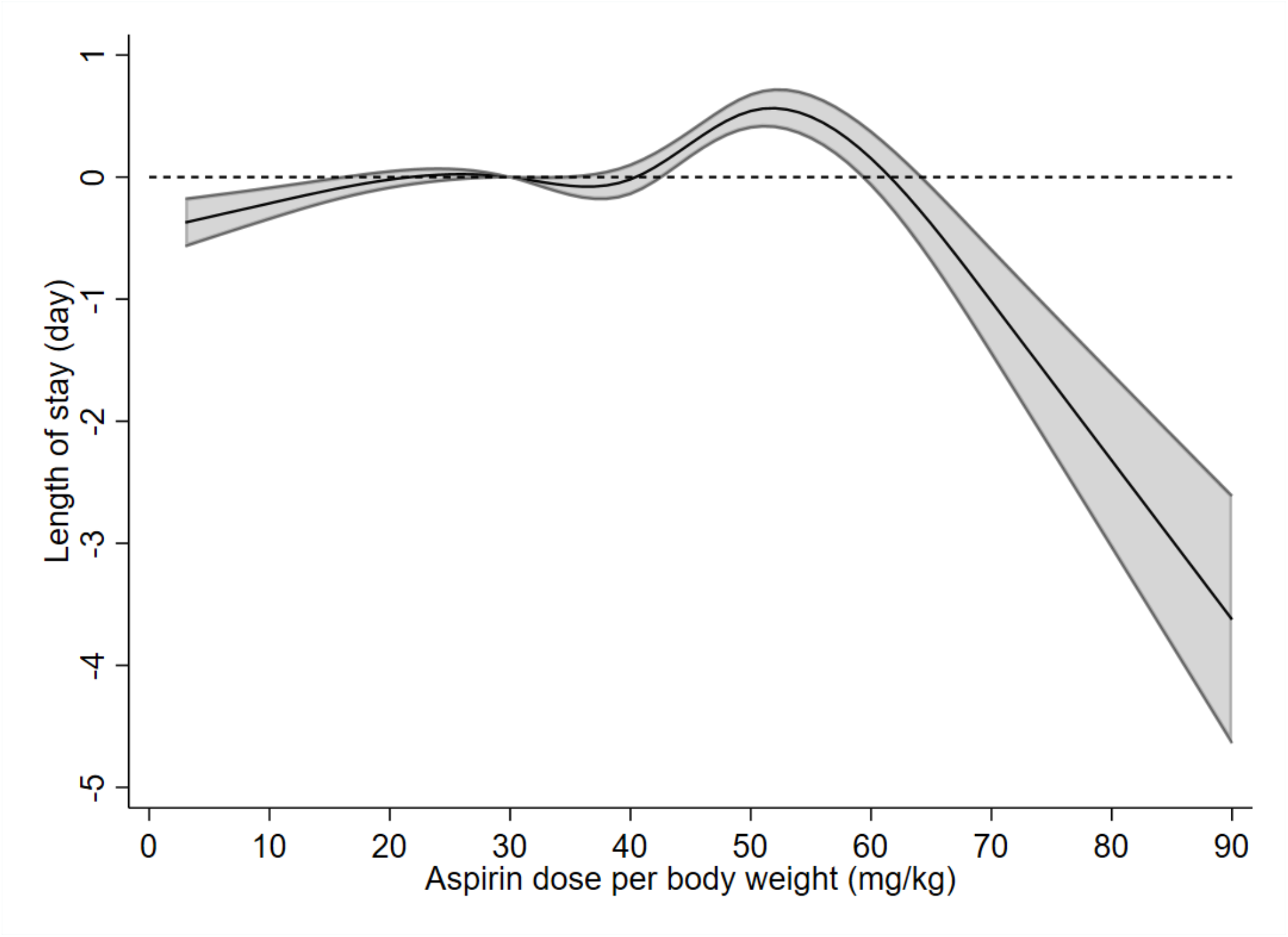
Restricted cubic spline for length of hospital stay The dose–response relationship between aspirin dose and the length of hospital stay was adjusted by analysis using a restricted cubic spline with five knots (Aspirin, 10, 30, 40, 50, and 70 mg/kg/day). The graphs show the estimated values along with 95% CI. IVIG: intravenous immunoglobulin; CI: confidence interval

**Figure 5.**
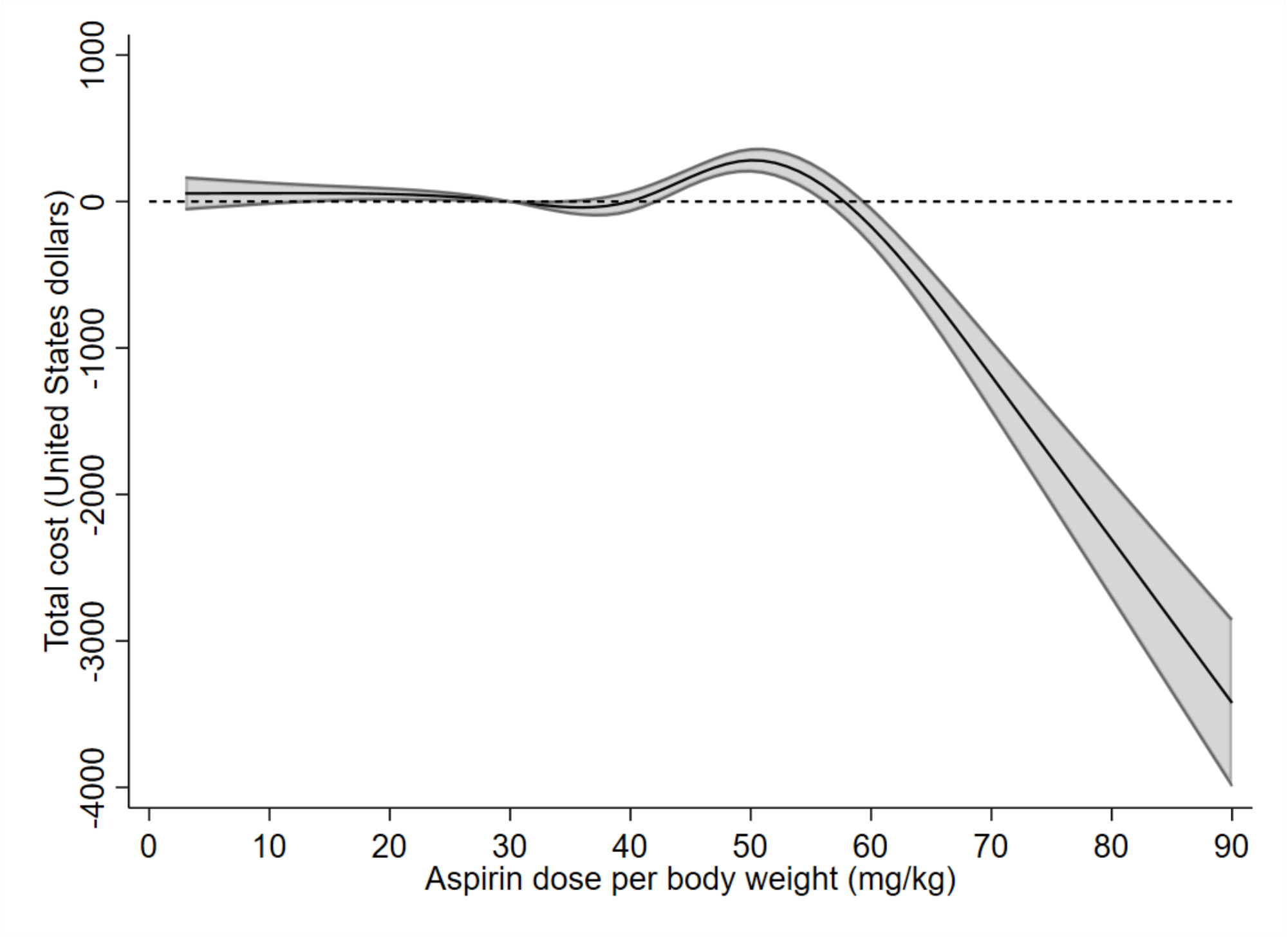
Restricted cubic spline for total medical cost The dose–response relationship between aspirin dose and the total medical cost was adjusted by analysis using a restricted cubic spline with five knots (Aspirin, 10, 30, 40, 50, and 70 mg/kg/day) The graphs show the estimated values along with 95% CI. IVIG: intravenous immunoglobulin; CI: confidence interval

In the sensitivity analysis for CAA, the odds ratio (95% CI) for CAA was 1.86 (1.33– 2.59) at 5 mg/kg/day compared with the values observed at 30 mg/kg/day. For aspirin doses of 30 mg/kg/day or higher, the 95% CIs for CAA were across 1 and not significantly different when compared with the values observed at 30 mg/kg/day (Figure 6).

**Figure 6.**
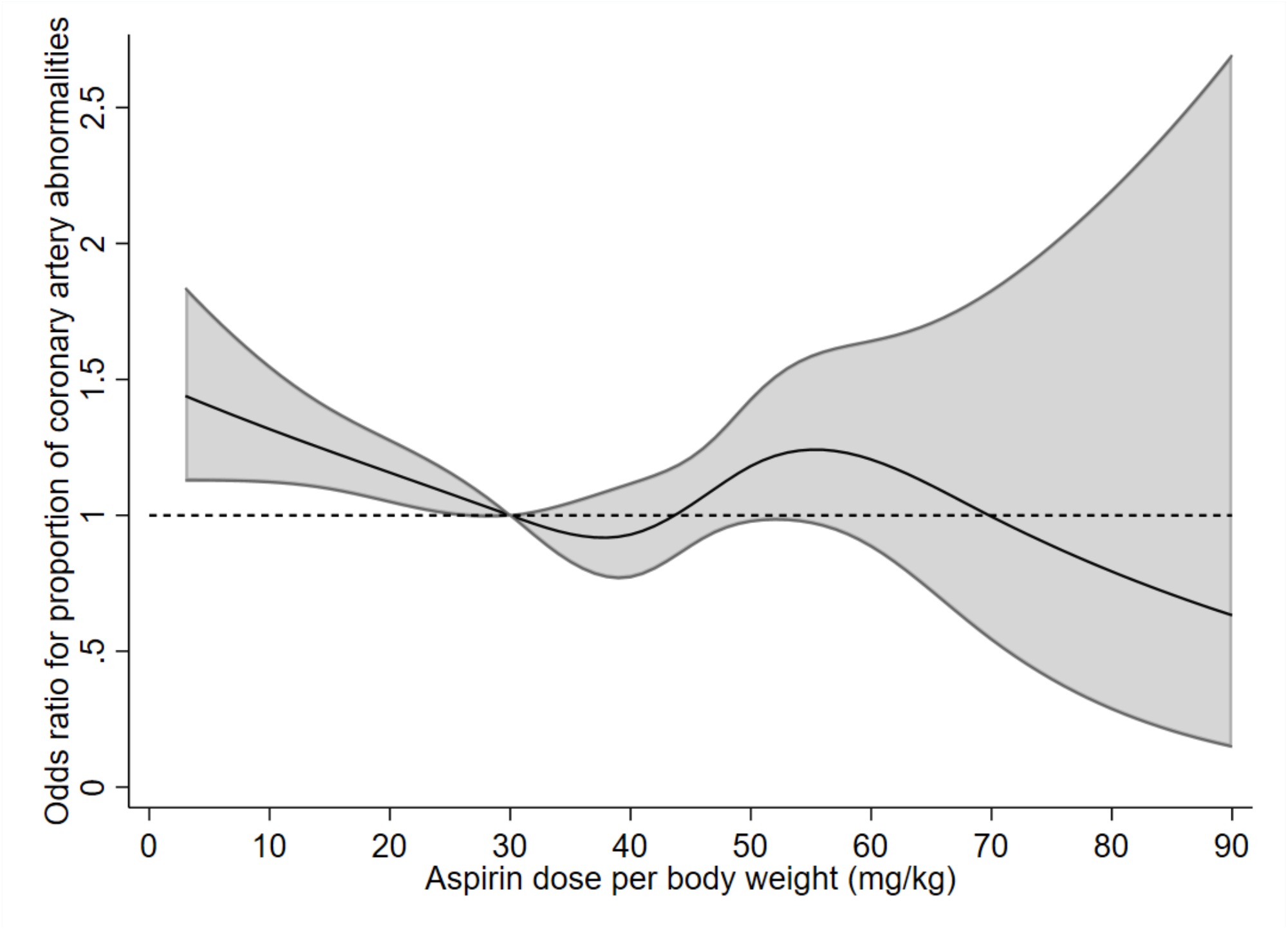
Restricted cubic spline for coronary artery abnormalities in the sensitivity analysis The dose–response relationship between aspirin dose and proportion of coronary artery abnormalities in the sensitivity analysis (limited to those with anticoagulant use or cardiac catheterization as well as diagnosis of coronary artery abnormalities) was adjusted by analysis using a restricted cubic spline with five knots (Aspirin, 10, 30, 40, 50 and 70 mg/kg/day). The graphs show the estimated values along with 95% CI. IVIG: intravenous immunoglobulin; CI: confidence interval

## Discussion

Our study evaluated serial aspirin doses from low (3 mg/kg/day) to high doses (90 mg/kg/day). Aspirin was generally considered to have antiplatelet effects at low doses and anti-inflammatory effects at standard or high doses. In this study, we examined a larger number of patients than those in previous studies and found that low-dose aspirin was significantly associated with a higher proportion of CAA. We also found that standard and high doses (over 50 mg/kg/day) of aspirin did not significantly change the occurrence of CAA.

The results of previous studies were inconsistent regarding the association between aspirin dose and CAA occurrence.^6,10,11,16^ Our results suggest that at least an anti-inflammatory dose of aspirin may be needed to reduce inflammation and prevent CAA in KD, and aspirin above the anti-inflammatory dose may neither suppress nor enhance the inflammatory response of patients with acute KD. Since these mechanisms are unknown, further basic research on KD is required to elucidate these mechanisms.

We showed that the low-dose and high-dose aspirin (>50 mg/kg/day) was associated with less proportion of IVIG resistance in patients with acute KD. Some previous reports indicated that lower aspirin doses may increase IVIG resistance,^7,8,16^ while others showed no significant difference in aspirin dose for IVIG resistance.^4-6,9^ The reason is unknown; however, clinicians may use low-dose aspirin in mild cases of patients with acute KD.

The present study showed that the use of high-dose aspirin (over 50 mg/kg/day) was associated with a shorter length of hospital stay and lower medical costs. Previous studies reported similar results for length of stay due to faster resolution of fever.^17,18^ Considering the results for length of stay and medical costs, the use of high-dose aspirin (over 50 mg/kg/day) may have resulted in early discharge from the hospital due to early resolution of fever, thus resulting in lower medical costs.

The present study examined possible adverse effects of aspirin and found no association between aspirin dose and adverse effects. A previous study showed anemia after IVIG treatment in patients who received over 30 mg/kg/day of aspirin.^5^ Another study showed gastrointestinal hemorrhage related to high-dose aspirin over 70 mg/kg/day.^19^ Many previous studies reported liver dysfunction due to KD inflammation, while liver dysfunction due to Reye’s syndrome was shown in patients with aspirin doses of 100 mg/kg/day.^20,21^ In our study, no cases of Reye syndrome were encountered. In general, side effects are often dose-dependent; but in this study, the proportion of side effects in the high-dose group (over 50 mg/kg/day) was not high.

## Limitations

We acknowledge several limitations in this study. First, our database lacked data regarding KD symptoms, laboratory findings, and fever duration of patients with acute KD. It is impossible to know from our database the days of illness during admission of the patient to the hospital. Therefore, we considered the days of hospital stay as days of illness. Patients who received steroids or cyclosporine are likely to have a higher severity of KD as per the Japanese Kawasaki disease score for severity classification.^22,23^ Second, the severity of CAAs due to KD, which was expressed as coronary arterial diameters or Z scores, was not included in our database. To clarify that CAAs were not transient, we used the history of cardiac catheterization and the prescription history of anticoagulants in addition to aspirin for sensitivity analysis. Third, adverse events due to aspirin were identified with recorded diagnoses, but these may have been underreported. Fourth, a few patients in our database used aspirin at doses greater than 90 mg/kg/day. Finally, because most of the patients in this study were Japanese and the generalizability of the present results to other populations may be limited.

## Data Availability

The datasets analyzed during the current study are not publicly available due to contracts with the hospitals providing data to the database.

## Conclusion

The results showed no significant association between aspirin escalation and CAAs in patients with acute KD. High-dose aspirin showed the potential to reduce hospital stay and medical costs without increasing complications.

## Abbreviations

ADL: activities of daily living
CAA: coronary artery abnormalities
CI: confidence interval
ICD-10: International Classification of Diseases, Tenth Revision
IQR: interquartile range
IVIG: intravenous immunoglobulin
KD: Kawasaki Disease
SD: standard deviation
USD: United States Dollar

## References

1. Newburger JW, Takahashi M, Burns JC. Kawasaki disease. J Am Coll Cardiol. 2016;67(14):1738–1749.

2. Mccrindle BW, Rowley AH, Newburger JW, et al. Diagnosis, treatment, and long-term management of Kawasaki disease: A scientific statement for health professionals from the American Heart Association. Circulation. 2017;135(17):e927–e999.

3. Miura M, Ayusawa M, Ito S. The guidelines on acute stage Kawasaki disease treatment. Pediatr Cardiol Card Surg. 2020;36(Suppl 1):S1.1–S1.29.

4. Hsieh KS, Weng KP, Lin CC, Huang TC, Lee CL, Huang SM. Treatment of acute Kawasaki disease: aspirin’s role in the febrile stage revisited. Pediatrics. 2004;114(6):e689–e693.

5. Kuo HC, Lo MH, Hsieh KS, Guo MMH, Huang YH. High-dose aspirin is associated with anemia and does not confer benefit to disease outcomes in Kawasaki disease. PLOS ONE, Woo PC, ed. 2015;10(12):e0144603.

6. Dallaire F, Fortier-Morissette Z, Blais S, et al. Aspirin dose and prevention of coronary abnormalities in Kawasaki disease. Pediatrics. 2017;139(6):e20170098.

7. Kim GB, Yu JJ, Yoon KL, et al. Medium-or higher-dose acetylsalicylic acid for acute Kawasaki disease and patient outcomes. J Pediatr. 2017;184:125–129.e1.

8. Zheng X, Yue P, Liu L, et al. Efficacy between low and high dose aspirin for the initial treatment of Kawasaki disease: current evidence based on a meta-analysis, editor. PLOS ONE. 2019;14(5):e0217274.

9. Jia X, Du X, Bie S, Li X, Bao Y, Jiang M. What dose of aspirin should be used in the initial treatment of Kawasaki disease? A meta-analysis. Rheumatology (Oxford). 2020;59(8):1826–1833.

10. Simon TD, Berry J, Feudtner C, et al. Children with complex chronic conditions in inpatient hospital settings in the United States. Pediatrics. 2010;126(4):647–655.

11. Yumoto T, Naito H, Yorifuji T, Aokage T, Fujisaki N, Nakao A. Association of Japan Coma Scale score on hospital arrival with in-hospital mortality among trauma patients. BMC Emerg Med. 2019;19(1):65.

12. Royston P, Altman DG, Sauerbrei W. Dichotomizing continuous predictors in multiple regression: a bad idea. Stat Med. 2006;25(1):127–141.

13. Desquilbet L, Mariotti F. Dose-response analyses using restricted cubic spline functions in public health research. Stat Med. 2010;29(9):1037–1057.

14. Ito Y, Matsui T, Abe K, et al. Aspirin dose and treatment outcomes in Kawasaki disease: A historical control study in Japan. Front Pediatr. 2020;8:249.

15. Nakada T. Effects of anti-inflammatory drugs on intravenous immunoglobulin therapy in the acute phase of Kawasaki disease. Pediatr Cardiol. 2015;36(2):335–339.

16. Lau AC, Duong TT, Ito S, Yeung RSM. Intravenous immunoglobulin and salicylate differentially modulate pathogenic processes leading to vascular damage in a model of Kawasaki disease. Arthritis Rheum. 2009;60(7):2131–2141.

17. Akagi T, Kato H, Inoue O, Sato N. A study on the optimal dose of aspirin therapy in Kawasaki disease—clinical evaluation and arachidonic acid metabolism. Kurume Med J. 1990;37(3):203–208.

18. Lee G, Lee SE, Hong YM, Sohn S. Is high-dose aspirin necessary in the acute phase of Kawasaki disease? Korean Circ J. 2013;43(3):182–186.

19. Matsubara T, Mason W, Kashani IA, Kligerman M, Burns JC. Gastrointestinal hemorrhage complicating aspirin therapy in acute Kawasaki disease. J Pediatr. 1996;128(5 Pt 1):701–703.

20. Wei CM, Chen HL, Lee PI, Chen CM, Ma CY, Hwu WL. Reye’s syndrome developing in an infant on treatment of Kawasaki syndrome. J Paediatr Child Health. 2005;41(5-6):303–304.

21. Lee JH, Hung HY, Huang FY. Kawasaki disease with Reye syndrome: report of one case. Zhonghua Min Guo Xiao Er Ke Yi Xue Hui Za Zhi. 1992;33(1):67–71.

22. Kobayashi T, Saji T, Otani T, et al. Efficacy of immunoglobulin plus prednisolone for prevention of coronary artery abnormalities in severe Kawasaki disease (RAISE study): a randomised, open-label, blinded-endpoints trial. Lancet. 2012;379(9826):1613–1620.

23. Hamada H, Suzuki H, Onouchi Y, et al. Efficacy of primary treatment with immunoglobulin plus ciclosporin for prevention of coronary artery abnormalities in patients with Kawasaki disease predicted to be at increased risk of non-response to intravenous immunoglobulin (KAICA): a randomised controlled, open-label, blinded-endpoints, phase 3 trial. Lancet. 2019;393(10176):1128–1137.

